# Sustained Oropouche virus transmission in Rio de Janeiro’s Atlantic forest: genomic evidence over a two-year period

**DOI:** 10.1101/2025.05.28.25328307

**Authors:** Ighor Arantes, Fernanda de Bruycker-Nogueira, Carla de Oliveira, Patrícia Carvalho Sequeira, Otília Lupi, Michele Borges, Carolina Lopes Melo, Michelle Brendolin, Antonio Braga, Luciane de Souza Velasque, Andréa Cony Cavalcanti, Adriana Cardoso Camargo, Fábio Burack da Costa, Cristiane Gomes de Castro Moreira, Vanessa Zaquieu Dias, Thiago de Jesus Sousa, Felipe Donateli Gatti, Gabriela Colombo de Mendonça, Joana Zorzal Nodari, Rodrigo Ribeiro-Rodrigues, Edson Delatorre, Guilherme Calvet, Felipe Gomes Naveca, Patricia Brasil, Ana Maria Bispo de Filippis, Gonzalo Bello, the OROV-Rio de Janeiro Outbreak Response Working Group

## Abstract

Since January 2024, over 1,500 Oropouche virus (OROV) cases have been confirmed in the Brazilian state of Rio de Janeiro. Genomic analyses showed that a lineage introduced in Rio de Janeiro in early 2024 persisted throughout 2025, highlighting the virus’s capacity for sustained transmission beyond the endemic Amazon region.

---

Oropouche virus (OROV) is an arbovirus belonging to the *Peribunyaviridae* family, genus *Orthobunyavirus*, that causes Oropouche fever, a neglected tropical disease, usually clinically indistinguishable from other arboviral diseases such as dengue, Zika, and chikungunya (1). In contrast to other urban arboviruses circulating in Latin America, OROV is endemic to the Amazon biome and is primarily transmitted to humans by the biting midge *Culicoides paraensis*, a vector adapted to both sylvatic and semi-urban environments (2). Between late 2022 and early 2024, four Brazilian states in the western Amazon region (Amazonas, Acre, Rondônia, and Roraima) experienced extensive OROV outbreaks, resulting in over 6,000 laboratory-confirmed cases linked to the spread of a newly reassortant viral lineage designated OROV_BR-2015-2024_ (3).

In 2024, the first confirmed OROV outbreaks outside the Brazilian Amazon were reported in several small municipalities within the Atlantic Forest biome (4). In early 2024, the OROV_BR-2015-2024_ lineage spread multiple times from the Amazon basin to non-endemic Atlantic Forest regions of Brazil, successfully establishing local transmission in Southern, Southeastern, and Northeastern states (5,6). Viral clades originating in the state of Amazonas, designated OROV_AM-I_ and OROV_AM-II_, seeded outbreaks in Santa Catarina (OROV_SC-I_) and Espírito Santo (OROV_ES-I_), respectively. Meanwhile, a third clade, OROV_AMACRO-II,_ circulating in the Northern states of Acre and Rondônia, gave rise to lineages detected in Santa Catarina (OROV_SC-II_), Espírito Santo (OROV_ES-II_ and OROV_ES-III_), and across both Rio de Janeiro and Espírito Santo (OROV_RJ/ES_).

The Brazilian state of Rio de Janeiro (**Figure 1a**) reported a small OROV outbreak in 2024 (n = 151), mainly in the Southern Fluminense region, followed by a increase in cases through April 30, 2025 (n = 1,075), particularly in rural areas of the Metropolitan and Northern Fluminense regions. Notably, most OROV cases were concentrated in small inland municipalities along Atlantic Forest areas (**Figure 1b**). Whether the 2025 epidemic in Rio de Janeiro resulted from a new viral introduction or ongoing transmission of the OROV_RJ/ES_ lineage introduced in 2024 remains unclear. We conducted genomic surveillance and phylodynamic analyses of OROV circulating in Rio de Janeiro state in 2025 to address (or to investigate) this question.

**Figure 1.**
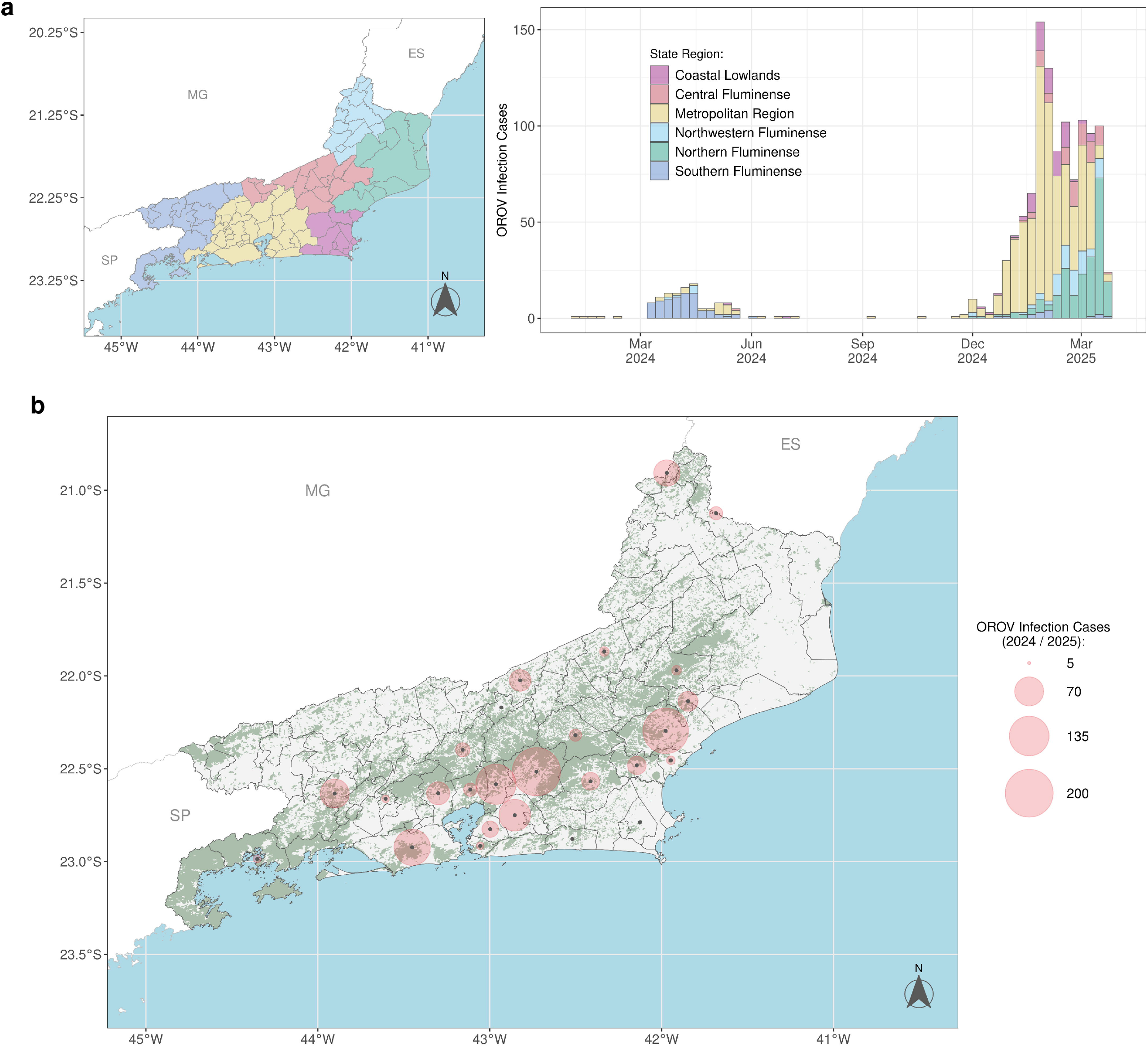
Spatial and temporal distribution of confirmed OROV cases in Rio de Janeiro state, 2024-2025. **a)** Temporal distribution of confirmed OROV cases, aggregated by epidemiological week (bars), and concurrently stratified by administrative region of infection within Rio de Janeiro state for the 2024-2025 period. Geographical regions depicted on the state map (left) are indicated by color (inset legend). **b)** Cumulative spatial distribution of confirmed OROV cases at the municipal level across the surveillance period. The total number of cases per municipality is represented proportionally by the diameter of the red circles; municipalities reporting five or more cases are explicitly marked. Areas shaded in green serve to delineate regions characterized by forest coverage, corresponding to official 2022 data from the Brazilian Institute of Geography and Statistics (IBGE, https://www.ibge.gov.br/), accessed via the R package geobr v.1.9.0. Abbreviations: ES: Espírito Santo; MG: Minas Gerais; SP: São Paulo. OROV cases were obtained from the Brazilian Ministry of Health’s official epidemiological dashboard (https://www.gov.br/saude/pt-br/assuntos/saude-de-a-a-z/o/oropouche/painel-epidemiologico) accessed on April 30, 2025.

## THE STUDY

This study was approved by the Ethics Committee of Instituto Oswaldo Cruz (CAAE: 90249218.6.1001.5248). Access to the genetic heritage of the OROV under investigation is registered in the National System for the Management of Genetic Heritage and Associated Traditional Knowledge (SisGen A0C0D2F). A total of 35 novel OROV genomes (complete or partial) were generated from cases detected in 15 municipalities across Rio de Janeiro state in 2024 (n = 16) and 2025 (n = 19) **(Figure 2a-b, Table S1)**. These cases were identified by RT-qPCR (7), as part of a collaborative surveillance effort involving the Rio de Janeiro state Central Laboratory, the Instituto Nacional de Infectologia Evandro Chagas from FIOCRUZ, and the Laboratorio de Arbovirus e Virus Hemorragicos at Instituto Oswaldo Cruz, FIOCRUZ, Rio de Janeiro, Brazil. Selected samples were submitted to total RNA extraction and used in sequencing library preparation following an adapted version of Illumina’s COVIDseq assay previously optimized for OROV sequencing (3). Consensus genomes were assembled using ViralFlow version 1.2.0 (8) and aligned with selected full or near-full-length OROV sequences (≥70% of coverage) with complete data for location and date of sampling (**Table S2**). Phylogenetic and phylogeographic analyses were performed using software IQ-TREE v.2.0.7 (9), Tempest v1.5.3 (10), and BEAST v1.10.4 (11) (**Appendix**). Consensus sequences are available at GISAID (https://doi.org/10.55876/gis8.250523gx).

**Figure 2.**
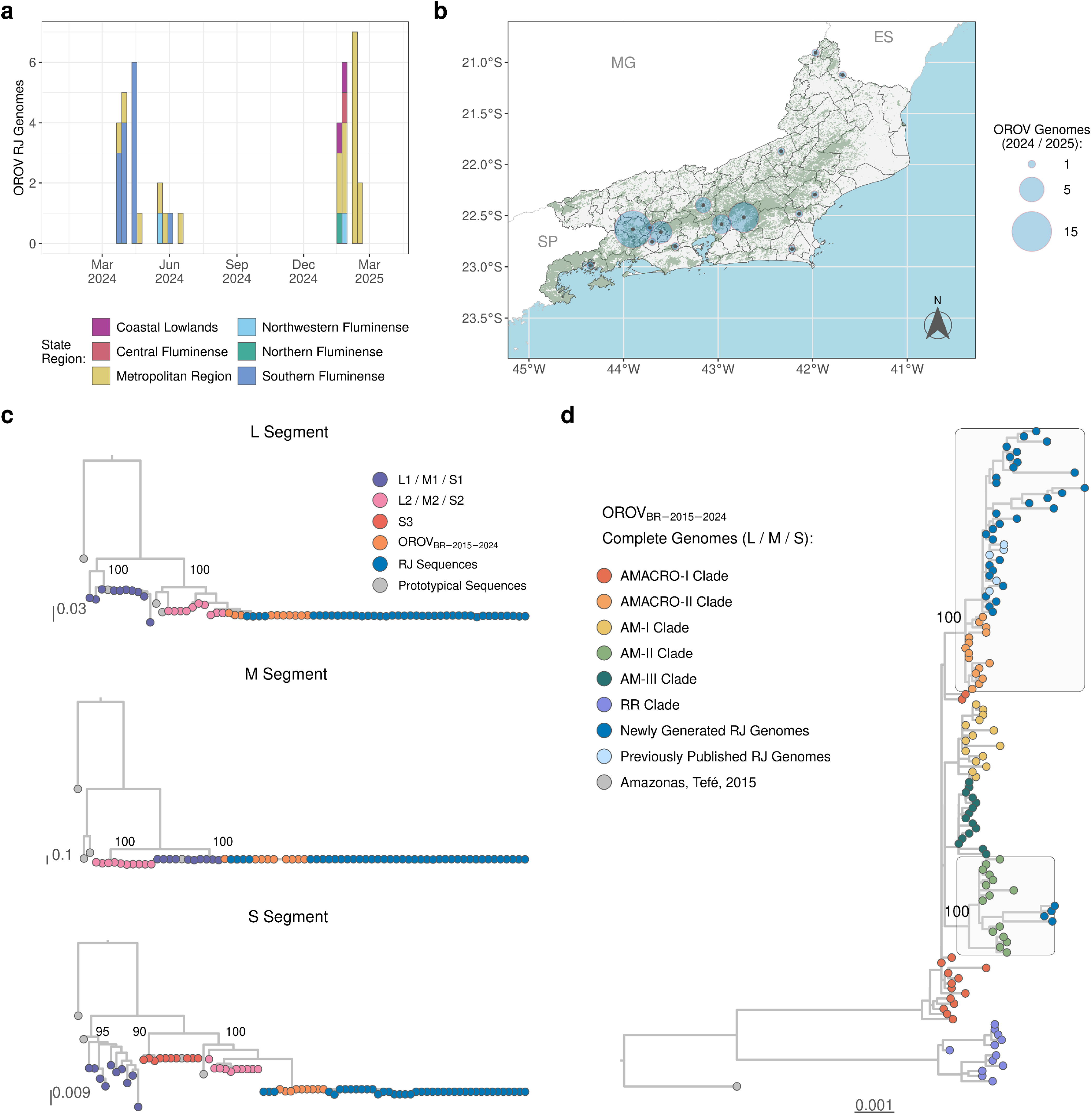
OROV genomes from Rio de Janeiro state, 2024-2025. **a)** Temporal distribution of OROV genomes from Rio de Janeiro (n = 40), aggregated by epidemiological week (represented by bars) for the 2024-2025 surveillance period and concurrently stratified by the administrative region where the infection likely occurred. **b)** Cumulative spatial distribution map displaying the locations of sampled OROV genomes at the municipal level throughout the surveillance period. The diameter of the blue circles represents the total number of genomes sequenced per municipality. Areas shaded in green indicate regions of forest coverage. **c)** Genotyping of OROV L, M, and S genome segments via ML phylogenetic trees. Sequences were colored according to their lineage/clade as indicated in the key located in the upper right corner. Sequences from Rio de Janeiro are indicated in blue and prototypical sequences (OROV, Perdões Virus, Madre de Dios Virus, and Iquitos Virus) in grey. Branch support values, calculated using the approximate Likelihood Ratio Test (aLRT), are annotated on key nodes defining the main lineages. **d)** ML phylogenetic tree of concatenated OROV segments. The dataset includes representatives of all clades previously identified within the OROV_BR-2019-2024_ lineage. Sub-clades containing genomes from Rio de Janeiro are highlighted and annotated with their statistical support values.

The Maximum likelihood (ML) phylogenetic analyses of individual genomic segments confirmed that all new genomes from Rio de Janeiro clustered within the OROV_BR-2015-2024_ lineage (**Figure 2c**). Next, a concatenated alignment of genomic segments L, M, and S was generated, comprising all OROV sequences from Rio de Janeiro and sequences representative of Amazonian sub-clades previously described (5). The new ML analysis indicates that most OROV sequences from Rio de Janeiro (n = 36) were nested within the OROV_AMACRO-II_ sub-clade, while four sequences branched within the OROV_AM-II_ sub-clade (**Figure 2d**). To model the OROV diffusion process in Rio de Janeiro, we thus included in subsequent analyses the earliest known sequences of the OROV_BR-2015-2024_ clade detected in 2015 and 2020, all OROV sequences from the Amazonian region classified within sub-clades OROV_AMACRO-I_, OROV_AMACRO-II_, and OROV_AM-II_, and all sequences from non-Amazonian states nested within those Amazonian sub-clades (3,5).

The correlation between genetic divergence and sampling time supports a significant (*p* < 0.05) temporal structure for the selected OROV dataset of concatenated genomic segments (**Figure 3a**). Bayesian discrete phylogeographic analysis revealed that all OROV sequences from Metropolitan, Southern Fluminense, and Northwest Fluminense regions grouped within the OROV_RJ/ES_ sub-clade. In contrast, those from the Coastal Lowlands, Northern Fluminense, and Central Fluminense regions belonged to the OROV_ES-I_ sub-clade (**Figure 3b**). The OROV_RJ/ES_ sub-clade likely originated in Acre state (*Posterior Sate Probability* [*PSP*] = 1) and was probably introduced into the Southern Fluminense region (*PSP* = 0.67) around 18 January 2024 (95% Highest Posterior Density [HPD]: 16 December 2023 - 13 February 2024). It subsequently spread to the Metropolitan region and from there to the Northwest Fluminense region and Espirito Santo, before moving back from Espirito Santo into the Northwest Fluminense region (**Figure 3c**). The OROV_ES-I_ sub-clade was introduced from Espírito Santo state into Rio de Janeiro’s Central Fluminense and Coastal Lowlands regions, subsequently spreading from the former to the Northern Fluminense region (**Figure 3d**).

**Figure 3.**
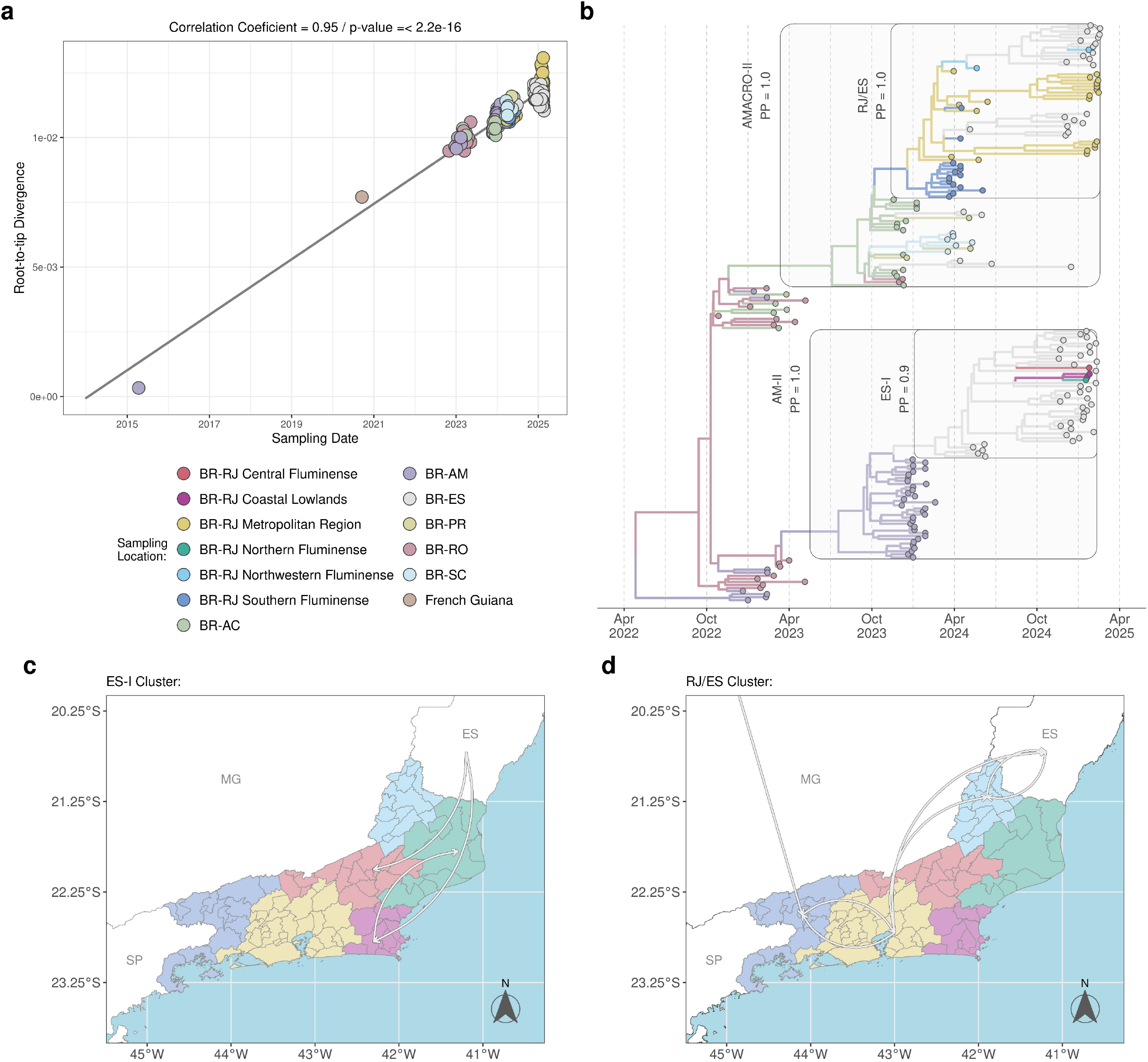
Discrete phylogeographic analysis of OROV dissemination in Rio de Janeiro state, 2024-2025. **a)** Evaluation of temporal signal through a linear regression of root-to-tip genetic divergence versus sampling date for selected OROV dataset (n = 191) comprising the oldest OROV_BR-2015-2024_ lineage genomes (Amazonas state, 2015; French Guiana, 2020) and the complete AMACRO-I (n = 28), AMACRO-II (n = 86), and AM-II (n = 75) clades. Data points are color-coded according to sampling location, corresponding to the legend at the panel’s base. **b)** Time-scaled Bayesian phylogeographic reconstruction inferred using the previously described genomic dataset. Branches are colored based on the inferred discrete ancestral location state, employing the color scheme from panel (a). Clades of interest are highlighted (light gray background) and annotated with established nomenclature and *Posterior Probability* (*PP*) support values. The basal 2015 and 2020 sequences were excluded from the graphical display for improved clarity. **c-d)** Schematic illustrations summarizing the inferred viral migration pathways involving Rio de Janeiro state, as deduced from the phylogeographic analysis presented in (b). Panel (c) delineates migration dynamics associated with the RJ/ES cluster, whereas panel (d) illustrates those pertinent to the ES-I cluster. Both maps employ the color scheme for state administrative regions defined in panel (a). Abbreviations: BR: Brazil; AC: Acre; AM: Amazonas; ES: Espírito Santo; PR: Paraná; RJ: Rio de Janeiro; RO: Rondônia; RR: Roraima; SC: Santa Catarina.

We applied a continuous spatial diffusion model with non-homogeneous dispersion rates to reconstruct the fine-scaled dispersion of the OROVRJ/ES sub-clade. Most viral migration events occurred over very-short (<2 km, 50%) and short (2–10 km, 30%) distances **(Figure 4a)**. The average dispersion rate of the OROV_RJ/ES_ sub-clade was estimated at 0.3 km/day (95% HPD: 0.2 - 0.4 km/day) **(Figure 4b)**, consistent with known active flight ranges of *Culicoides spp*. vectors (12–14). From January 2024 to January 2025, the median OROV dispersion rate dropped from 0.5 km/day to 0.1 km/day **(Figure 4c)**. The epicenter of the OROV_RJ/ES_ sub-clade was placed in the municipality of Piraí, in the Southern Fluminense region, from where the virus moved northwards into the Metropolitan and Northwest Fluminense regions in early 2024 (**Figure 4d**). The OROV_RJ/ES_ sub-clade persisted undetected in the Metropolitan region during the second half of 2024 and seeded new outbreaks in early 2025.

**Figure 4.**
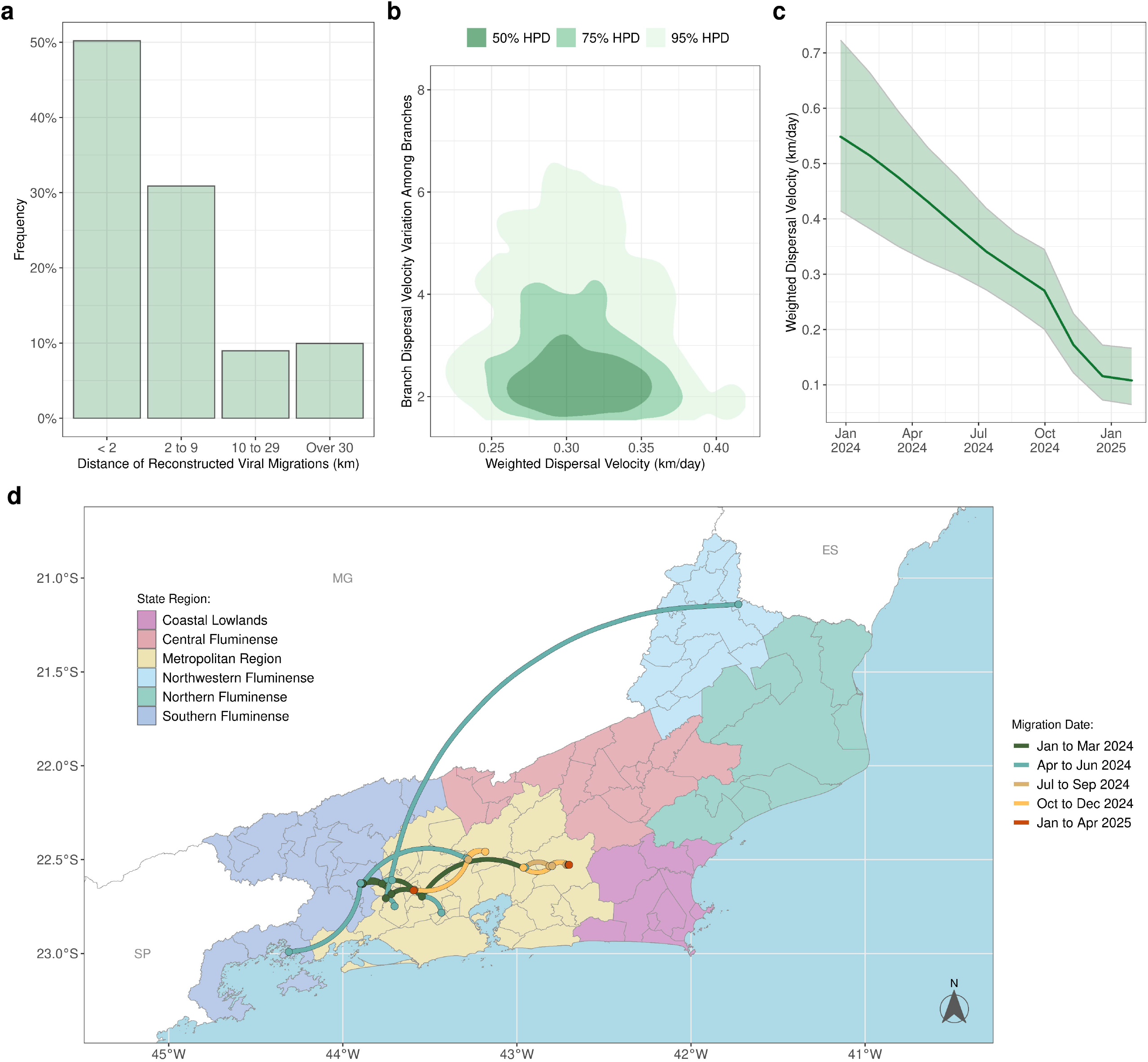
Continuous phylogeographic analysis of OROV dissemination in Rio de Janeiro state, 2024-2025. The panels present results inferred via continuous phylogeographic reconstruction, focusing on OROV genomes from Rio de Janeiro belonging to the RJ/ES cluster (n = 35). **a)** Distribution of inferred intra-state migration distances, stratified by distance category: very short (< 2 km), short (2 – 9 km), medium (10 – 29 km), and large (≥ 30 km). **b)** Joint posterior density estimate (represented by a 2D kernel density plot) for the weighted dispersal velocity (km/day, x-axis) versus the among-branch variation in this velocity (y-axis). Shaded areas denote the 50% (dark gray), 75% (medium gray), and 95% (light gray) HPD intervals for the joint parameter estimates. **c)** Temporal evolution trajectory of the estimated weighted dispersal velocity (km/day). The thicker line indicates the median posterior estimate, with the corresponding 95% HPD interval represented by the light green shaded area. **d)** Spatial projection of the inferred viral dissemination pathways among the Rio de Janeiro regions. Each curved line represents a corresponding branch from the Bayesian Maximum Clade Credibility tree. Lines are depicted as curved trajectories with a clockwise orientation. The color of each line segment corresponds to the estimated mean date of that lineage’s occurrence, according to the color key on the panel’s left side.

## CONCLUSIONS

Our findings confirm that the 2025 OROV epidemic in the state of Rio de Janeiro was primarily driven by sustained local transmission of the OROV_RJ/ES_ sub-clade, introduced in early 2024, along with multiple independent introductions from the neighboring state of Espírito Santo. Our study supports that the OROV_RJ/ES_ sub-clade primarily spreads among contiguous inland municipalities located near forested areas in Rio de Janeiro, likely facilitated by the dispersal capabilities of local vectors. These results highlight the potential for sustained autochthonous OROV transmission across multiple years in Atlantic Forest regions of Rio de Janeiro state, and point to the risk of extra-Amazonian spread of OROV within the Southeastern region of Brazil. These findings suggest that the OROV may establish endemic circulation beyond the Amazon region and underscore the importance of routinely including OROV in differential diagnoses of acute febrile illnesses in the Southeastern Brazilian region.

### Members of the OROV-Rio de Janeiro Outbreak Response Working Group

Cintia Damasceno dos Santos Rodrigues, Carolina Cardoso dos Santos, Juan Carlos Proença Moura, Desiree dos Santos Nunes, Simone Alves Sampaio (Laboratório de Arbovírus e Vírus Hemorrágicos, Instituto Oswaldo Cruz, FIOCRUZ, Rio de Janeiro, Brazil); Claudia Maria Braga, Mario Sergio Ribeiro, Debora Fontenelle dos Santos, Silvia Cristina de Carvalho Cardoso, Cristina Giordano, Paula Almeida, (Secretaria de Estado de Saúde do Rio de Janeiro); Lusiele Guaraldo, Luana Damasceno, Thais Pires Trindade, Leticia Lopes Corrêa, Fernanda Moronoe, Heloisa Ferreira, Trevon Fuller, Isabella Moraes, Marise Mattos, Ezequias Batista Martins, Rogério Valls, Anielle Pina Costa, Roxana Flores, Clarisse Bressan, Manuela da Costa Medeiros, Stephanie Penetra, Otávio de Melo Espíndola, (Laboratório de Doenças Febris Agudas, Instituto Nacional de Infectologia Evandro Chagas, FIOCRUZ, Rio de Janeiro, Brazil); Ieda Pereira Ribeiro, Myrna Cristina Bonaldo (Laboratório de Medicina Experimental e Saúde - IOC/Fiocruz - RJ). Elizabeth Portari (Instituto Fernandes Figueira, Fiocruz, RJ).

## Data Availability

All OROV Consensus sequences are available at GISAID.

https://doi.org/10.55876/gis8.250523gx

## Acknowledgments

The authors would like to thank Tânia M Peixoto Fonseca from the Coordination of Health Surveillance and Reference Laboratories (CVSLR)/Fiocruz for the financial support, to the Next Generation Sequencing Platform (RPT01J) - Technology Platforms Network/VPPCB - FIOCRUZ, and the Respiratory, Exanthematous, Enterovirus and Viral Emergencies Laboratory of the Oswaldo Cruz Institute (IOC/Fiocruz) for their support in building and reading the libraries, and to collaborators Solange Regina da Conceição and Ronaldo Lapa Lopes for their technical support.

## Authors contributions

I.A., F.B-N., G.C., F.G.N., P.B., A.M.B.dF. and G.B. contributed to the conception and design of this work. P.B. and A.M.B.dF. obtained ethical approval. F.B-N., C.O., P.C.S, O.L., M.B., C.L.M., M.B., A.B., L.dS.V., A.C.C., A.C.C., G.C., and P.B. contributed to diagnostics, patient recruitment, sample collection, and public health surveillance data analysis from Rio de Janeiro state. F.B-N. and C.O. contributed with genomic sequencing and raw sequencing data analysis. T.dJ.S., F.D.G., G.C.dM., J.Z.N., R.R-R., E.D, F.B.C, C.G.C.M and V.Z.D contributed to diagnostics, public health surveillance data and genomic data from Espirito Santo state. I.A., F.G.N. and G.B. contributed to phylogenetic data analysis. F.G.N., P.B, and A.M.B.dF. contribute with funding acquisition and project administration. I.A and G.B wrote the original draft with the support of F.B-N., P.C.S., G.C., F.G.N., P.B., and A.M.B.dF. All authors had substantial contributions to the acquisition and interpretation of the data and approved the final version of the manuscript.

## About the Authors

Dr. Arantes and Dr. Bruycker-Nogueira are public health researchers at Brazil’s Fundação Oswaldo Cruz, working in Rio de Janeiro. Their primary research interest lies in diagnostics, molecular epidemiology, and the evolution of emergent and reemergent viruses.

