## Appendix for "Sustained Oropouche virus transmission in Rio de Janeiro’s Atlantic forest: genomic evidence over a two-year period"

### SUPPLEMENTARY TABLES

**Table S1**: Novel complete Oropouche virus genomes sequenced from Rio de Janeiro State.

| ID | Accession ID | Ct | Municipality | Sampling Year | Origin |
|---|---|---|---|---|---|
| 4816_2024 | EPI_ISL_19880158 | 20.89 | Piraí | 2024 | LACEN/RJ |
| 4827_2024 | EPI_ISL_19880159 | 17.97 | Piraí | 2024 | LACEN/RJ |
| 5216_2024 | EPI_ISL_19880160 | 25.54 | Japeri | 2024 | LACEN/RJ |
| 5323_2024 | EPI_ISL_19880161 | 21.12 | Piraí | 2024 | LACEN/RJ |
| 5844_2024 | EPI_ISL_19880163 | 19.19 | Seropédica | 2024 | LACEN/RJ |
| 5396_2024 | EPI_ISL_19880162 | 20.76 | Piraí | 2024 | LACEN/RJ |
| 6305_2024 | EPI_ISL_19880164 | 24.2 | Guapimirim | 2024 | LACEN/RJ |
| 6408_2024 | EPI_ISL_19880165 | 28.36/28.5 | Piraí | 2024 | LACEN/RJ |
| 6412_2024 | EPI_ISL_19880166 | 29.63/29.57 | Piraí | 2024 | LACEN/RJ |
| 6413_2024 | EPI_ISL_19880167 | 27.6/27.47 | Piraí | 2024 | LACEN/RJ |
| 6676_2024 | EPI_ISL_19880168 | 20.06 | Piraí | 2024 | LACEN/RJ |
| 8161_2024 | EPI_ISL_19880156 | 25.82/25.97 | Bom Jesus do Itabapoana | 2024 | LACEN/RJ |
| 8162_2024 | EPI_ISL_19880157 | 23.32/23.36 | Mesquita | 2024 | LACEN/RJ |
| 8158_2024 | EPI_ISL_19880153 | 28.21/28.56 | Paracambi | 2024 | LACEN/RJ |
| 8159_2024 | EPI_ISL_19880154 | 29.23/29.46 | Angra dos Reis | 2024 | LACEN/RJ |
| 8160_2024 | EPI_ISL_19880155 | 24.48/24.54 | Guapimirim | 2024 | LACEN/RJ |
| 0849_2025 | EPI_ISL_19880020 | 21.86/20.68 | Macaé | 2025 | LACEN/RJ |
| 0854_2025 | EPI_ISL_19880139 | 21.69/21.76 | Petrópolis | 2025 | LACEN/RJ |
| 0855_2025 | EPI_ISL_19880140 | 23.66/23.68 | Iguaba Grande | 2025 | LACEN/RJ |
| 0856_2025 | EPI_ISL_19880141 | 20.51/20.65 | Petrópolis | 2025 | LACEN/RJ |
| 0851_2025 | EPI_ISL_19880136 | 20.16/20.19 | Porciúncula | 2025 | LACEN/RJ |
| 0853_2025 | EPI_ISL_19880138 | 21.19/21.21 | Cachoeiras de Macacu | 2025 | LACEN/RJ |
| 0852_2025 | EPI_ISL_19880137 | 23.49/23.38 | Cantagalo | 2025 | LACEN/RJ |
| 0857_2025 | EPI_ISL_19880142 | 22.16/22.49 | Casimiro de Abreu | 2025 | LACEN/RJ |
| 0858_2025 | EPI_ISL_19880143 | 21.88/21.99 | Guapimirim | 2025 | LACEN/RJ |
| 0850_2025 | EPI_ISL_19880135 | 20.09/19.19 | Cachoeiras de Macacu | 2025 | LACEN/RJ |
| 0883_2025 | EPI_ISL_19880144 | 21.13/21.05 | Japeri | 2025 | INI |
| 0886_2025 | EPI_ISL_19880145 | 16.91/16.83 | Japeri | 2025 | INI |
| 1209_2025 | EPI_ISL_19880147 | 21.76/22.03 | Cachoeiras de Macacu | 2025 | INI |
| 1217_2025 | EPI_ISL_19880148 | 20.9/20.95 | Cachoeiras de Macacu | 2025 | INI |
| 1221_2025 | EPI_ISL_19880149 | 19.39/19.62 | Cachoeiras de Macacu | 2025 | INI |
| 1226_2025 | EPI_ISL_19880150 | 19.31/19.18 | Cachoeiras de Macacu | 2025 | INI |

| | | | | | |
|---|---|---|---|---|---|
| 1201_2025 | EPI_ISL_19880151 | 21.95/22.25 | Japeri | 2025 | INI |
| 1241_2025 | EPI_ISL_19880151 | 30.14/30.2 | Cachoeiras de Macacu | 2025 | INI |
| 1273_2025 | EPI_ISL_19880152 | 27.62/27.73 | Cachoeiras de Macacu | 2025 | INI |

**Table S2**: Oropouche virus complete genomes of the $OROV_{BR-2015-2024}$ clade previously published.

| Database | From | To | Sampling Location | DOI |
|---|---|---|---|---|
| GISAID (https://gisaid.org/) | EPI_ISL_19706611 | EPI_ISL_19706627 | ES | NA |
| | EPI_ISL_19723915 | EPI_ISL_19723929 | | |
| | EPI_ISL_19810547 | EPI_ISL_19810568 | | |
| GeneBank (https://www.ncbi.nlm.nih.gov/genbank/) | PP153945 | PP154172 | AM, AC, RO, RR | 10.1038/s41591-024-03300 |
| | PQ064571 | PQ065491. | | |
| | PQ073181 | PQ073186 | ES, PE, PR, RJ, SC | 10.1016/S1473-3099(24)00687-X |
| | PQ156583 | PQ156627 | | |
| | PQ189413 | PQ189445 | | |
| | PQ295361 | PQ295375 | | |
| | OL689332 | OL689334 | GF | 10.3201/eid 2710.204760 |

Abbreviations: AC: Acre; AM: Amazonas; BR: Brazil; GF: French Guiana; NA: Not Available; PE: Pernambuco; PR: Paraná; RO: Rondônia; RR: Roraima; SC: Santa Catarina.

### MATERIALS AND METHODS

**OROV-positive Samples and ethics.** This study was approved by the Ethics Committee of Instituto Oswaldo Cruz (CAAE: 90249218.6.1001.5248). Access to the genetic heritage of the OROV under investigation is registered in the National System for the Management of Genetic Heritage and Associated Traditional Knowledge (SisGen A0C0D2F). As part of a collaborative surveillance effort involving the Rio de Janeiro state Central Laboratory (LACEN/RJ) and the Instituto Nacional de Infectologia Evandro Chagas (INI) from FIOCRUZ, 35 OROV-positive samples identified by RT-qPCR, with a cycle threshold (Ct) lower than 30 (26 samples received from LACEN/RJ and 9 from INI/Fiocruz), at the Arbovirus and Hemorrhagic Virus Laboratory of the Oswaldo Cruz Institute, FIOCRUZ, Rio de Janeiro, Brazil, were submitted to viral genomic sequencing.

**Whole-genome sequencing and genome assembling.** OROV-positive samples were submitted to total RNA extraction using the Thermo Scientific™ KingFisher™ Flex Purification System and used in sequencing library preparation following an adapted version of Illumina's COVIDseq assay previously optimized for OROV sequencing (2). Library preparation was carried out using Illumina's COVIDSeq kit, and sequencing was performed on a MiniSeq version 2.3.0 instrument with the Mid-Output Reagent Cartridge kit (300 cycles) for a 151 bp × 2 paired-end run. FASTQ reads were generated on Illumina BaseSpace (https://basespace.illumina.com), and consensus genomes were assembled using ViralFlow version 1.2.0 (3). Reference sequences AF484424, AF441119, and AY237111 from GenBank were used for the L, M and S, virus segments of OROV, respectively.

**OROV Whole-genome Genotyping.** The complete (>70% of coverage) OROV consensus sequences of the S, M, and L genomic segments from Rio de Janeiro, both generated in this study (*n* = 35) and previously published (*n* = 5), were genotyped following the method described by Naveca *et al*. (2). All RJ genomes were aligned using MAFFT v7.505 (4) against curated reference datasets specific to each designated OROV genome segment. The datasets included (I) a subset of all genomes previously assigned to each segment type (L1/L2, M1/M2, S1/S2/S3) (2) clusterized with CD-HIT v.2.4.8 (5); (II) an also clustered subset of all $OROV_{BR-2015-2024}$ ($L_2M_1S_2$) clade genomes available until March 2024; and (III) the following prototype virus sequences, all classified within the species *Orthobunyavirus oropoucheense*: OROV (GenBank accessions L: AF484424, M: AF441119, S: AY237111), Iquitos virus (L: KF697142, M: KF697143, S: KF697144), Perdões virus (L: KP691627, M: KP691628, S: KP691629), and Madre de Dios virus (L: KF697147, M: KF697145, S: KF697146). The datasets were used to infer maximum likelihood (ML) trees for each OROV genomic segment using IQ-TREE v2.2.2.7 (6) under the best nucleotide substitution model selected by the ModelFinder software (7). Branch support was assessed using the approximate likelihood-ratio test (aLRT) (8) based on the Shimodaira–Hasegawa-like procedure with 1,000 replicates. The ML trees were visualized using Figtree v.1.4.4 (9), and the Treeio v3.1.7 (10), and the ggtree v3.2.1 (11) R packages. Genotypes were defined by the clustering of RJ sequences with reference sequences exhibiting robust statistical support (aLRT > 0.90).

**$OROV_{BR-2015-2024}$ Subclade Assignment**. All OROV genomes from Rio de Janeiro state previously assigned to the $OROV_{BR-2015-2024}$ clade were further resolved into its subclades (2). To construct a dataset for this analysis, representative sequences from established subclades (AMACRO-I, AMACRO-II, AM-I, AM-II, AM-III and RR-I) were selected by a clusterization approach using CD-HIT v.2.4.8 (5). This dataset was supplemented with the oldest known genome from this lineage, sampled in Tefé, Amazonas, (2015). A ML phylogenetic tree was then inferred using the parameters previously described. The assignment of RJ sequences to specific established clades was determined by the formation of a monophyletic group with reference sequences from that clade, supported by robust statistical evidence (aLRT > 0.90).

**Bayesian Discrete Phylogeographic Analysis.** To elucidate the introduction pathways and subsequent interstate dissemination dynamics of OROV in Rio de Janeiro state, a comprehensive phylogeographic dataset was assembled. This dataset comprised: (I) all available OROV sequences originating from Rio de Janeiro state; (II) all available sequences from the parental clades of these Rio de Janeiro sequences, located in the Amazonian regions; and (III) the two chronologically earliest genomes identified within the $OROV_{BR-2015-2024}$ subclade, which were sampled in Tefé, Amazonas (2015), and French Guiana (2020). The temporal structure of this selected dataset was evaluated using TempEst v1.5.3 (12) through a root-to-tip linear regression. The significance of the correlation between collection date and genetic distance was assessed using a Pearson correlation test, with adjustments for multiple comparisons made via the Benjamini–Hochberg method for false discovery rate control. Time-scaled phylogenetic trees were estimated using the MCMC approach implemented in BEAST v1.10.4 (13), with BEAGLE v4.0.0 (14) employed to improve computational efficiency. Bayesian trees were reconstructed under the nucleotide substitution model selected by the ModelFinder application (7), the non-parametric Bayesian Skyline coalescent demographic model (15), and a relaxed molecular clock model with a continuous-time Markov chain (CTMC) rate reference prior (16). The time to the most recent common ancestor ($T_{MRCA}$), and the spatial dynamics of the identified RJ clusters were reconstructed using a discrete phylogeographic model (17) implemented in BEAST v1.10.4 (13). For this analysis, we employed a CTMC rate reference prior and a symmetric phylogeographic model. Reflecting the focus on reconstructing the introduction and dissemination dynamics of OROV within Rio de Janeiro state, sequences originating from this state were assigned to their most probable region of infection; in contrast, all other sequences (i.e., those not from Rio de Janeiro state) were coded either by their Brazilian state of sampling or, if international, by their country of origin. Adequate mixing and convergence of the Markov Chain Monte Carlo (MCMC) run were confirmed by ensuring that all continuous parameters achieved an effective sample size (ESS) exceeding 200, as verified using Tracer v1.7 (12). The maximum clade credibility (MCC) tree was summarized with TreeAnnotator v1.10 (13) and visualized using using Figtree v.1.4.4 (9) and also the Treeio v3.1.7 (10) and ggtree v3.2.1 (11) R packages. The spatial dissemination patterns of major Oropouche virus (OROV) clusters across the administrative regions of Rio de Janeiro state were geographically visualized on maps. generated using the ggplot2 (17), sf v.1.0-20 (18), and geobr v.1.9.1 (19) R packages.

**Bayesian Continuous Phylogeographic Analysis.** The intrastate OROV dissemination dynamics in Rio de Janeiro and its diffusion rate were additionally inferred using a continuous phylogeographic analysis employing a heterogeneous relaxed random walk model with a Cauchy distribution (20). For this analysis, an inclusion criterion was established whereby only clusters comprising more than 30 genomes sampled within Rio de Janeiro state were selected. This threshold was implemented to ensure the robust estimation of cluster-specific epidemiological parameters associated with viral dissemination and to more accurately capture the dynamics of viral circulation within the state. Sequences were coded based on the spatial coordinates of their city of origin, and for sequences from the same municipality, a noise factor (0.01 degrees) was added to the sampling coordinates to ensure each sequence had unique geographic coordinates. The weighted diffusion rate and its variation across branches was inferred using the seraphim R package (21), and migratory events were summarized with the cross-platform software SPREAD v1.0.7 (22). The spatiotemporal diffusion patterns of the virus and its associated epidemiological parameters were, respectively, projected onto maps and graphically plotted. These visualizations and analyses were conducted in R, utilizing the packages geobr v.1.9.1 (19), ggplot2 (17), sf v.1.0-20 (18), geosphere v.1.5-18 (23) and HDInterval v.0.2.4 (24).

### References


1. Naveca FG, Nascimento VA do, Souza VC de, Nunes BTD, Rodrigues DSG, Vasconcelos

PF da C. Multiplexed reverse transcription real-time polymerase chain reaction for simultaneous detection of Mayaro, Oropouche, and Oropouche-like viruses. Mem Inst Oswaldo Cruz. 2017 Jul;112(7):510–3.
